# Lack of Accuracy of the GRACE score to Predict Coronary Anatomy in Acute Coronary Syndromes

**DOI:** 10.1101/2021.01.26.21250301

**Authors:** Mateus dos Santos Viana, Thomaz Emanoel Azevedo Silva, Gabriela Oliveira Bagano, Bruna de Sá Barreto Pontes, Milton Henrique Vitoria de Melo, Pedro Henrique Correia Filgueiras, Andre Costa Meireles, Paula Oliveira de Andrade Lopes, Andre Luiz Freitas de Oliveira Junior, João Vitor Miranda de Oliveira Porto, Vitor Calixto de Almeida Correia, Luis Claudio Lemos Correia

## Abstract

**Introduction:** Coronary anatomy is one of the strongest risk predictors in Acute Coronary Syndromes (ACS), which justifies early coronary angiography. Diagnostic scores for predicting outcomes are usually superior to clinical judgment. Despite being validated for prognosis, the GRACE score has been used to discriminate patients with high or low probability of anatomical severity.

**Objective:** To test the hypothesis that the GRACE score actually predicts anatomical severity.

**Methods:** The study was carried out by assessing consecutive patients with ACS who underwent invasive angiography. Severe anatomical disease was defined as obstructive involvement (≥ 70% in diameter) in (1) left main coronary artery or (2) double or triple vessel disease involving proximal left anterior descending artery or (3) subocclusion. The GRACE score was evaluated under numerical and dichotomous tests.

**Results:** A total of 733 patients were evaluated, aged 63 ± 14 years, 61% male and GRACE score of 119 ± 37. Obstructive coronary disease was observed in 81% of the patients, classified as one, two or three vessel disease, or left main coronary artery involvement in 28%, 23%, 26% and 4%, respectively. The area under the ROC curve of the GRACE score was 0.65 (95% CI = 0.61 - 0.69) for predicting severe disease. The cutoff point below which the first GRACE tertile is defined (109) was used to dichotomize low-risk (N = 318) and medium-high-risk (N = 415) samples. This standard definition of intermediate-high risk by the GRACE score (> 109) revealed sensitivity of 67% in detecting severe anatomy (95% CI = 61% - 72%) and specificity of 50% (95% CI = 46% - 55%), resulting in positive likelihood ratio of 1.3 (95% CI = 1.2 - 1.5) and negative likelihood ratio of 0.66 (95% CI = 0.55 - 0.80). There was a weak correlation between GRACE and anatomical scores such as SYNTAX (r = 0.36, P < 0.001) and Gensini (r = 0.36, P < 0.001).

**Conclusion:** Despite statistical association with extent of anatomical coronary disease, the GRACE Score is not accurate to predict severity of disease before coronary angiography.

## INTRODUCTION

The prognosis of acute coronary syndromes (ACS) varies according to clinical and anatomical characteristics^1^. Probabilistic risk scores are used to assess prognosis based on clinical data, and their results usually guide the selection of patients better suited for invasive strategy, in which coronary anatomy is assessed early. However, clinical risk scores are not well established as predictors of coronary disease’s anatomical severity.

Among existing models, the GRACE score is the most accurate for prognosis, being a reliable predictor of death during hospitalization and after 6 months of discharge in patients with ACS^2^. Previous studies indicated positive correlation between the GRACE score and angiographic scores^3,4^, although a comprehensive analysis of its accuracy for severe coronary anatomy had not been carried out before.

The present study aims beyond correlation and evaluates the accuracy of the GRACE score regarding the severity of ACS.

## METHODS

### Research design

This is a cross-sectional diagnostic study, derived from prospective registry of ACS.

### Sample selection

All patients admitted to the coronary care unit with a diagnosis of ACS from July 2007 to December 2017 were included in a Registry of Acute Coronary Syndromes. The criteria for inclusion was defined as typical chest discomfort at rest in the last 48 hours, associated with at least one of the following: positive myocardial necrosis marker, defined by troponin T ≥ 0.01 µg/L or troponin I > 0.034 µg/L, which correspond to values above the 99 percentile^5,6^, ischemic electrocardiographic changes, consisting in T wave inversion (≥ 0.1 mV) or transient ST segment depression (≥ 0.05 mV) or coronary artery disease previously documented, defined by a myocardial infarction history or previous angiography showing coronary obstruction ≥ 50%. For the present analysis, only patients who underwent invasive coronary angiography during hospitalization were selected. Exclusion criteria were previous revascularization surgery or a patient’s refusal to participate in study.

The protocol is in accordance with the Declaration of Helsinki^7^, was approved by the Institution’s Research Ethics Committee and all evaluated patients signed informed consent forms.

### Anatomical definitions

Obstructive coronary artery disease was defined as stenosis ≥ 70% in any epicardial coronary artery or obstruction ≥ 50% in left main coronary artery. Severe anatomical disease was defined as obstructive involvement (≥ 70% in diameter) in (1) left main coronary artery or (2) double or triple vessel disease involving proximal left anterior descending artery or (3) obstruction ≥ 95% in any proximal arterial segment.

All patients underwent a coronary anatomy analysis by an experienced interventional cardiologist, blind to treatment modality and clinical condition, who measured the number of vascular territories, categorized severe anatomy and quantified coronary disease extent by using different angiographic scores: modified Gensini, Friesinger, proximal disease score, Duke Jeopardy and SYNTAX.

Gensini score^8^ assesses 28 coronary segments, according to CASS map^9,10^, which is scored according to their anatomical relevance (ranging from 0.5 to 5) multiplied by a factor relating to maximum obstruction degree in diameter (0 - 25% = 2; 26% - 50% = 4; 51% −75% = 8; 76% - 90% = 16; 91% - 99% = 32; 100% = 64 points).

Friesinger score^11^ uses a simpler sum of 0 to 15, from three main coronary arteries being quantified from 0 to 5 (0 - without angiographic abnormalities; 1 - diameter irregularities with obstruction < 29%; 2 - localized obstruction with 30 - 68%; 3 - multiple or diffuse obstructions of 30 - 68% in diameter; 4 - luminal obstruction between 69 - 100%, without proximal involvement and 5 - total proximal segment obstruction).

Proximal disease score^8^ ranges from 1 to 7 points, with vessels with diameter reduction ≥ 70% being graded as: 1 - not compromised proximal segment; 2 - 1 proximal right coronary artery segment or circumflex artery affected; 3 – proximal anterior descending artery segment affected; 4 - both proximal involvement of right and circumflex coronary arteries; 5 – proximal anterior descending artery involvement and one of right or circumflex coronary arteries; 6 – Any left coronary trunk segment involvement; 7 – proximal three vessels involvement, with or without left main coronary artery involvement.

Duke Jeopardy^12^ angiographic score assesses vessels with > 75% obstruction in diameter, using a previously described^13^ six-segment coronary grading system to estimate myocardial mass at risk. Establishing two points for each segment, the total can vary from 0 to 12 points.

Lastly, SYNTAX^14^ score, which assesses each coronary with a diameter ≥ 1.5 mm and obstruction ≥ 50% following a previously described tutorial^14^, considering angiographic parameters such as location of lesion and number of affected vessels, presence of bifurcation lesion in coronary ostia, total artery occlusion, time of occlusion, collateral circulation, lesion extent, presence of thrombi, significant tortuosity, excessive calcification and diffuse disease.

### GRACE Score calculation

The GRACE score was calculated using clinical data from patients’ presentation in the emergency department, such as electrocardiographic records in the first 6 hours of hospital, troponins measurements in the first 12 hours of care and first serum creatinine. Myocardial marker of necrosis was defined as troponin T ≥ 0.01 µg/L or troponin I ≥ 0.034 µg/L, that is, above the 99 percentile^5,6^.

In summary, the GRACE score consists of eight variables: five of them are semi-quantitative, i.e. different weights for each age group, systolic blood pressure, heart rate, plasma creatinine and Killip class; three of them are dichotomous (ST segment depression, myocardial necrosis marker elevation, cardiac arrest at admission). The final score can vary from 0 to 372^15^, being the cutoff < 109 for low risk and ≥ 109 for medium/high risk.

### Statistical Analysis

The association of GRACE with anatomical severity was assessed by Spearman’s and Pearson’s correlation tests, according to normal distribution of the scores. Linear regression was used to demonstrate the influence of GRACE score in coronary disease extent. The Kruskal-Wallis test was used to compare score values between the groups divided according to GRACE tertiles.

The predictive accuracy of risk scores concerning the presence of CAD or severe anatomy was tested with the receiver-operating characteristics (ROC) curve. Sensitivity, specificity, positive and negative likelihood ratios and positive and negative predictive values were also described. The sample size required for correlation analysis was estimated considering a minimum correlation coefficient of 0.25 and an alpha of 0.05. A sample of 96 patients would be enough to provide 80% statistical power in rejecting the null hypothesis of r < 0.25. All patients consecutively present in the registry who met inclusion criteria were selected, with no voluntary choice of individuals, making a sample size much larger than estimated.

Statistical analyses were carried out using SPSS (Statistical Package for Social Sciences v. 21.1) and WinPepi (Copyright J.H. Abramson, August 23, 2016; version 11.65).

## RESULTS

### Sample Characteristics

Out of a total of 1103 patients included in the Registry, 975 patients (88%) underwent invasive coronary angiography. From these, 101 patients who had had previous myocardial revascularization surgery and 141 patients whose diagnostic test had not been properly recorded for analysis were excluded. The study sample therefore consisted of 733 patients aged 63 ± 14 years, 61% male and 33% with history of CAD (Table 1). Non-ST elevation acute myocardial infarction (50%) was the most common ACS presentation, followed by unstable angina (26%) and ST-elevation myocardial infarction (24%). The GRACE score has a median of 115 (IIQ 92 – 140). As outcomes, 3% of individuals had recurrent non-fatal infarction during hospitalization, and 4% had cardiovascular death (Table 2). Obstructive coronary disease was present in 81% of the patients, and 41% presented severe anatomy, with similar 1, 2, or 3 - vessel disease proportion, (28%, 23% and 26%, respectively). The median of SYNTAX score was 10 (IIQ 4 - 21) and for the Gensini score it was 114 (IIQ 72 - 170) (Table 3).

**Table 1.** Clinical characteristics of the sample.

| Characteristic | N |
| --- | --- |
| Sample size | 733 |
| Age (years) | 63 ± 14 |
| Male | 446 (61%) |
| BMI (kg/m<sup>2</sup>) | 27 ± 7 |
| Systemic arterial hypertension | 546 (75%) |
| Diabetes mellitus | 252 (34%) |
| Dyslipidemia | 444 (61%) |
| Current smoking | 82 (11%) |
| Family history of CAD | 245 (33%) |
| Previous CAD | 242 (33%) |
| Previous CVA | 57 (8%) |
| Previous infarction | 151 (21%) |
| Previous peripheral vascular disease | 41 (6%) |
| ACS presentation |  |
| STEMI | 180 (24%) |
| NSTEMI | 364 (50%) |
| Unstable angina | 189 (26%) |
| GRACE score | 119 ± 37 |
| Non-fatal infarction | 21 (3%) |
| Cardiovascular death | 31 (4%) |

**BMI** – body mass index, **CAD** – coronary artery disease, **CVA** – cerebrovascular accident, **ACS** – acute coronary syndromes, **STEMI** – ST-elevation myocardial infarction, **NSTEMI** – non-ST elevation myocardial infarction, **GRACE** – global registry of acute coronary events.

**Table 2.** Laboratorial characteristics and diagnostic tests of the sample.

| Characteristic | N |
| --- | --- |
| Sample size | 733 |
| Ischemic changes in ECG | 342 (47%) |
| Positive troponin | 422 (58%) |
| Killip classification > 1 | 102 (14%) |
| Serum level of Pro-BNP | 401 (115 – 1235) |
| Serum creatinine on admission (mg/dL) | 0.90 (0.80 – 1.10) |
| Clearance of creatinine (ml/min) | 88 (63 – 116) |
| Hemoglobin on admission (g/dL) | 13.8 ± 1.7 |
| Blood glucose on admission (mg/dL) | 120 (99 – 183) |
| LDL – cholesterol (mg/dL) | 105 ± 41 |
| HDL – cholesterol (mg/dL) | 41 ± 12 |
| Triglycerides (mg/dL) | 155 (90 – 178) |

*ECG* – electrocardiogram at rest, *BNP* – brain natriuretic peptide, *LDL* – low density lipoprotein, *HDL* – high density lipoprotein.

**Table 3.** Coronary anatomy pattern of the sample.

| Anatomic pattern | N |
| --- | --- |
| Compromised vascular territories |  |
| No significant obstructions | 138 (19%) |
| Uniarterials | 203 (28%) |
| Biarterials | 165 (23%) |
| Triarterial pattern | 191 (26%) |
| Left coronary trunk obstruction | 29 (4%) |
| Two or three proximal segments compromised | 102 (14%) |
| Proximal Arterial Segments score | 1 (1 – 3) |
| Duke Jeopardy score | 5.2 ± 3.8 |
| Friesinger score | 7.6 ± 3.9 |
| Modified Gensini score | 114 (72 – 170) |
| SYNTAX score | 10 (4 – 21) |

### Correlation of GRACE score with anatomical severity of disease

A weak correlation between the GRACE score and the SYNTAX score (r = 0.36; P < 0.001) or Gensini (r = 0.36; P < 0.001) was found, with an intercept of - 1.85 and a regression coefficient of 0.12 for SYNTAX and intercept values of 37 and regression coefficient of 0.79 for Gensini (Figure 1). Other anatomical variables show the same strength of association, as shown in Table 4.

**Table 4.** Correlation between GRACE score and angiographic data.

| Anatomic score | Inclination ($\beta$) | Intercept ($\alpha$) | Correlation (r) | P |
| --- | --- | --- | --- | --- |
| Number of arteries | 0.008 | 0.73 | 0.26 | < 0.001 |
| Number of proximal segments | 0.007 | - 0.30 | 0.35 | < 0.001 |
| Proximal Arterial Segments score | 0.015 | 0.34 | 0.33* | < 0.001 |
| Duke Jeopardy score | 0.034 | 1.17 | 0.33 | < 0.001 |
| Friesinger score | 0.034 | 3.56 | 0.33 | < 0.001 |
| Modified Gensini score | 0.780 | 38.04 | 0.36* | < 0.001 |
| SYNTAX score | 0.12 | - 1.52 | 0.36* | < 0.001 |

\* *Spearman's correlation coefficient was calculated for variables with non-normal distribution. Other coefficients were obtained using Pearson's test.*

**Table 5.** Accuracy of GRACE score to predict severe anatomy.

| Criterion | Severe anatomy (CI 95%) |
| --- | --- |
| Sensitivity | 67% (61% – 72%) |
| Specificity | 50% (46% – 55%) |
| Positive likelihood ratio | 1.3 (1.2 – 1.5) |
| Negative likelihood ratio | 0.66 (0.55 – 0.80) |
| Positive predictive value | 48% (45% – 52%) |
| Negative predictive value | 68% (64% – 72%) |

**Figure 1.**
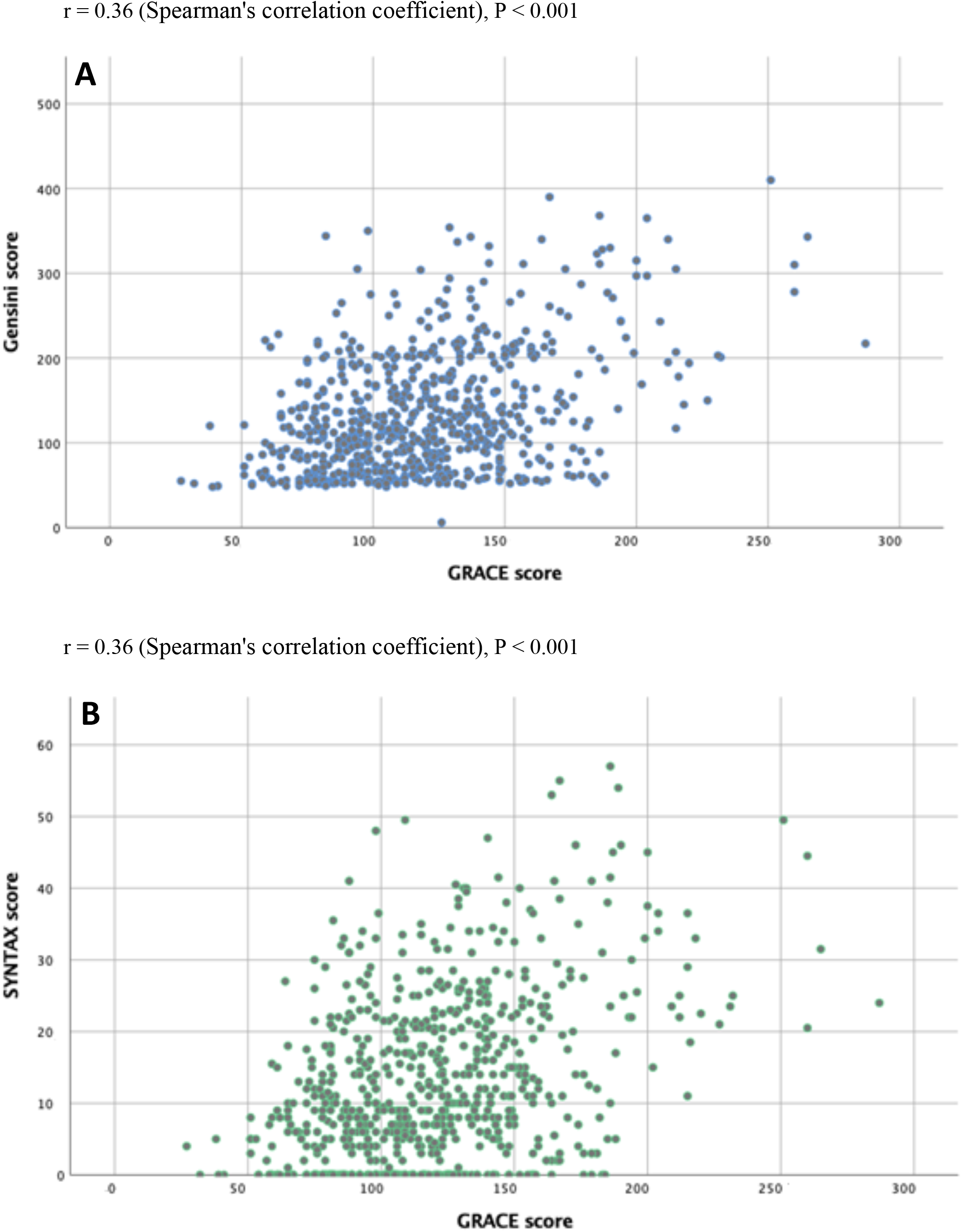
Correlation of GRACE score with angiographic severity according to Gensini (Panel A) and SYNTAX (Panel B) scores.

Accordingly, the extent of coronary disease increased with tertiles of the GRACE score in a moderate fashion. The SYNTAX score had medians of 7 (IIQ 0 - 14), 12 (IIQ 6 - 21) and 17 (IIQ 7 - 27), in the first, second and third tertiles (Figure 2A) respectively (P < 0.001), while the Gensini score had medians of 97 (IIQ 62 - 135), 122 (IIQ 85 - 174) and 122 (IIQ 85 - 174) (Figure 2B), respectively (P < 0.001).

**Figure 2.**
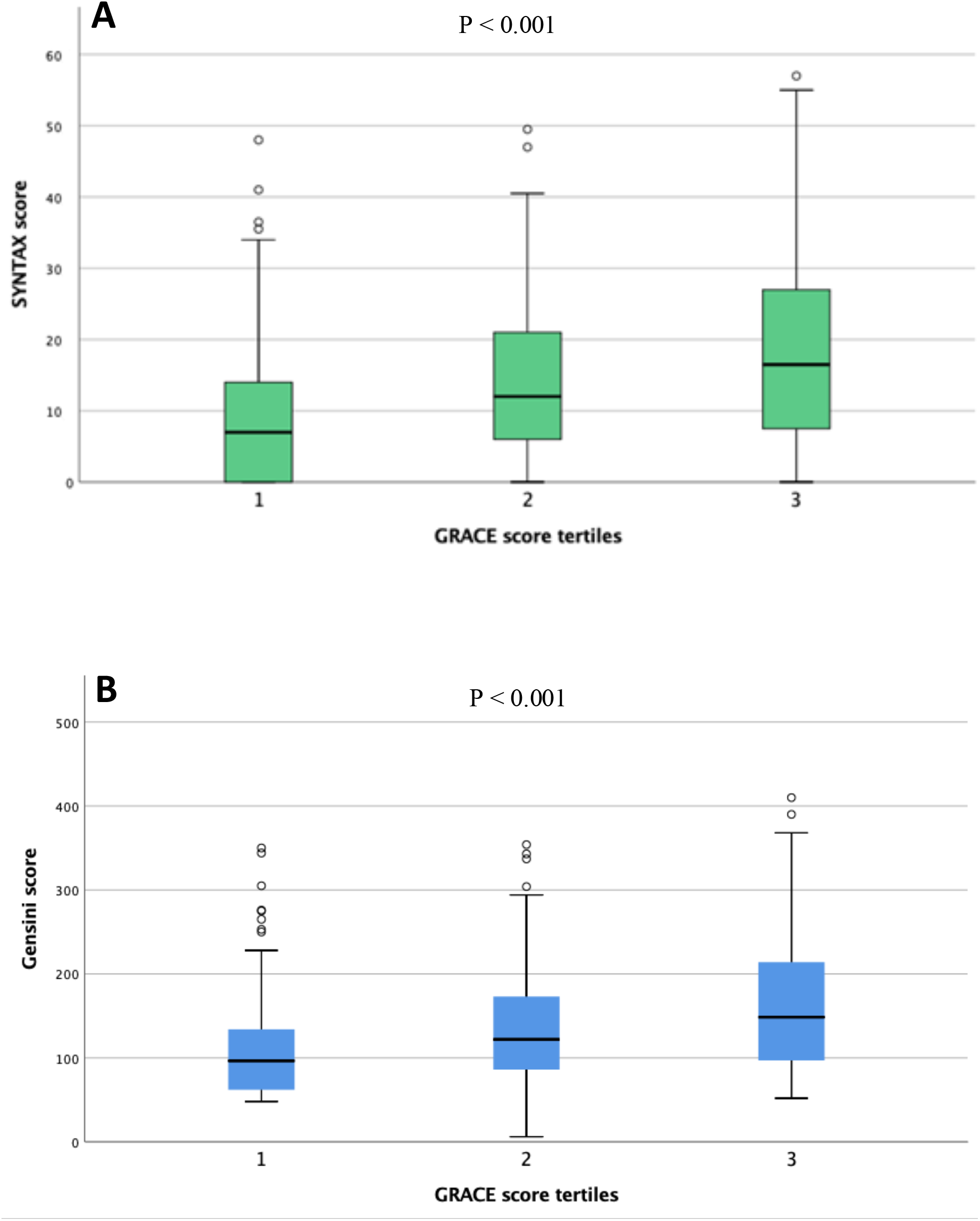
Comparison of angiographic severity in ACS between the respective tertiles of GRACE score according to SYNTAX (Panel A) and Gensini (Panel B) scores.

### Accuracy of the GRACE score for anatomical severity of CAD

The GRACE score has a modest discriminatory capacity in detecting severe anatomy, with an area under the ROC curve (Figure 3) of 0.65 (95% CI 0.60 - 0.69; P < 0.001). The prevalence of severe anatomy according to tertiles of this score was respectively 32%, 41% and 59% (P < 0.001).

**Figure 3.**
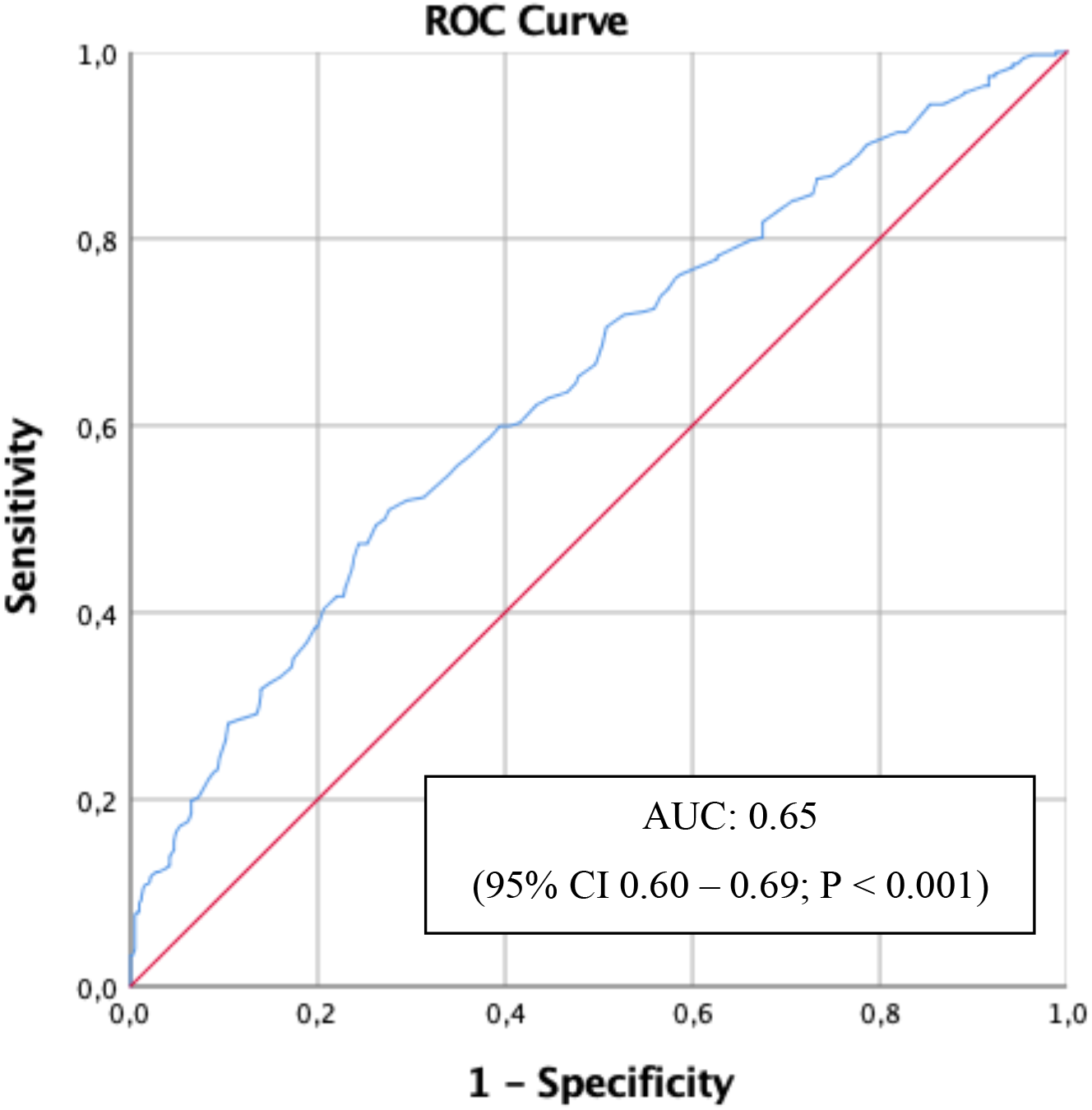
Predictive accuracy of GRACE score for severe anatomy in ACS.

Accuracy of traditional GRACE’s cut-off points proved to be weak for predicting severe anatomy. The cut-off point below which the first tertile of GRACE score (109) is defined was used to dichotomize the sample at low risk (N = 318) and medium-high risk (N = 415). This criteria showed sensitivity of 67% to detect severe anatomy (95% CI = 61% - 72%) and specificity of 50% to identify patients without severe anatomy (95% CI = 46% - 55%), resulting in a positive likelihood ratio of 1.3 (95% CI = 1.2 - 1.5) and a negative likelihood ratio of 0.66 (95% CI = 0.55 - 0.80), which were insufficient to satisfactorily increase or decrease the pretest probability of severe anatomy. Accordingly, the positive predictive value of GRACE ≥ 109 was 48% (95% CI = 45% - 52%), and the negative predictive value of GRACE < 109 was 68% (95% CI = 64% - 72%) for severe anatomy.

## DISCUSSION

This study indicates that the accuracy of the GRACE score for anatomical severity of CAD is modest. Coronary anatomy was analyzed both quantitatively, expressed in the form of angiographic scores, and qualitatively, defined as severe anatomy. In the former analysis, despite a linear association between angiographic and the GRACE scores, the correlation coefficient was weak. In accordance, there was a small area under the curve of GRACE to predict severe anatomy.

Previous studies^3,4^ concluded in favor of GRACE’s ability to predict coronary anatomy, but they solely based this conclusion on correlation analysis. This predictive value of the GRACE score was contested in a previous publication of preliminary findings. The present study confirms those preliminary findings^16^ with much greater depth, based on a sample seven times larger and more detailed coronary anatomy analyses, in addition to having determined the ROC curve, sensitivity, specificity and likelihood ratios and a categorization in risk groups using the traditional cut-off points of the score^2^.

The GRACE scoring system is based on multivariate models that integrate elements from medical history, admission ECG and biochemical myocardial necrosis evidence. This score is not intended to identify coronary artery disease extent and complexity, but its use as a predictor to determine the need for invasive stratification by coronary angiography is widespread, possibly based on a false premise that association equals accuracy.

A possible explanation for the modest accuracy of GRACE for anatomic prediction comes from the fact that anatomical disease extent represents only one of many severity determinants in a complex multivariate model, typical of most biological systems^16^. The GRACE model was generated and validated to predict mortality, but this outcome depends, mostly, on an individual’s susceptibility to ACS (age, fragility, comorbidities) and not only on ischemia burden. Secondly, the manifestation of myocardial infarction severity, clinically expressed by Killip’s classification, serum troponin levels, ECG wall extent and echocardiogram among other clinical data, are more dependent on variables such as time from onset of symptoms to reperfusion, reperfusion success and extent of myocardium affected by the artery related to the event than on atherosclerotic disease burden per se^17-23^.

A further remarkable aspect of this study is a linear association tested with a large number of angiographic scores, all pointing to the same direction. It is widely accepted that a positive correlation is not enough to ensure satisfactory accuracy. This should be evaluated by measures that identify unhealthy and healthy individuals, such as sensitivity and specificity, respectively. Furthermore, accuracy involves the concepts of positive and negative likelihood ratios, which in turn increases or decreases the pretest probability of a certain condition such as severe coronary anatomy. The results confirm a positive association of the GRACE score with coronary disease extent but demonstrate no satisfactory accuracy to predict such outcome.

A limitation of the study, to be covered in further research, was a selection bias for patients who underwent coronary angiography, to the exclusion of patients with lower risk or probable lesser disease extent.

## CONCLUSION

Based on the largest sample and set of data analyzed so far, this study indicated that the GRACE score is not an accurate tool to satisfactorily predict the extent of coronary disease in patients with ACS. The utility of this score in clinical reasoning should be limited to estimate risk in prognostic evaluations.

## Data Availability

All data were properly stored, under confidentiality and are are available upon reasonable request. Data may be obtained from a third party and are not publicly available.

